# Prevalence and correlates of age at first birth and utilization of maternal health care services among pregnant women in four Asian-Pacific Island countries

**DOI:** 10.1101/2023.10.05.23296593

**Authors:** Wah W. Myint, David J. Washburn, Shinduk Lee, Brian Colwell, Petronella Ahenda, Jay E. Maddock

**Author notes:** Corresponding Author. (wwm). Authors contribution (WWM contribute to conceptualization, data curation, formal analysis, funding acquisition, methodology, validation, writing – original draft, writing reviewed and revised. DJW, BC, and JEM equally contributed to the manuscript’s conceptualization, methodology, supervision, validation, writing – review & editing. SL contributed to conceptualization, methodology, validation, writing – review & editing. PA contributed to validation, writing – review & editing).

## Abstract

**BACKGROUND:** Maternal mortality is still high among adolescent women worldwide. This study explores the influence of socio-demographic characteristics on having four or more antenatal care (ANC4+) visits and use of Skilled Birth Attendants (SBA) among women in four island countries: Indonesia, the Philippines, Papua New Guinea, and Timor-Leste.

**METHODS:** Data from Demographic and Health Surveys of Indonesia, the Philippines, Papua New Guinea, and Timor-Leste were used. We studied the relationship of utilization of ANC4+ visits and SBA among women aged 15-49 years with socio-demographic variables including women’s age at first birth, age at first cohabitation, age at last birth, age groups, place of residence, education, wealth, birth order, number of living children, household ownership of a vehicle, and reproductive health knowledge. Descriptive, bivariate, and multivariable logistic regression analyses were performed.

**RESULTS:** Women at first birth before age 18 ranged from 18% in Indonesia to 31% in Papua New Guinea. In all countries, wealth status and having knowledge on modern contraceptives were significant predictors for having ANC4+ visits and SBA use. In Indonesia, women older than 25 were 1.38 times (95% CI=1.09-1.74) more likely to use SBA than those younger than 19. Each country demonstrated different patterns of ANC4+ visits and use of SBA that varied by education, place of residence, birth order, number of living children, and household ownership of a vehicle.

**CONCLUSION:** Having a high number of adolescent births highlighted a need for reproductive health knowledge reach to the young women, which can achieve through ANC4+ visits and use of SBA.

## Introduction

Maternal mortality is a major concern in low- and lower-middle-income countries, with the estimated global Maternal Mortality Ratio (MMR) remaining at about 211 per 100,000 live births (1). 75% of maternal mortality is due to complications from pregnancy and childbirth, such as postpartum bleeding, infections, and pregnancy-induced hypertension (1). In addition, maternal age at first birth younger than 18 poses an increased risk for complications (2).

The Sustainable Development Goals have a variety of targets (3), including targets to reduce MMR. Target 3.1 is to have less than 70 MMR per 100,000 live births by 2030 (3). The World Health Organization (WHO) also set five coverage maternal health related targets in 2021, of which two of them are utilization of four or more antenatal care (ANC4+) visits by 90% of pregnant women and to have 90% of births to use a Skilled Birth Attendant (SBA) by 2025 (4, 5). Garenne et al. (1997) found that lack of ANC visits increased the odds of maternal mortality almost 16 times more than those who had ANC visits (6). All SBAs (medical doctors, nurses, and midwives) play important roles as they are trained to assess life-threatening conditions (7). Compared to higher-income island countries in the region, such as Singapore and New Zealand, which had MMR of less than 10 per 100,000 live births in 2017, the lower-income countries such as Indonesia, the Philippines, Papua New Guinea, and Timor-Leste still have higher MMR at 273, 109, 145, and 142 per 100,000 live births respectively in 2018 (8). These island countries show a greater diversity of culture and geography which influence the age at first birth and pregnant women’s access to health care services. For example, Timor-Leste is extremely mountainous, while the others have numerous islands that make health care access difficult for many pregnant women (9, 10). Although SBA is common in the high and upper-middle-income countries in the Asia and the Pacific region, they are scarce in these countries (Organization for Economic Co-operation and Development (11).

This study’s overall objective was to explore the relationships between the age at first birth and the use of maternal health services (i.e., use of ANC4+ and SBA) among women in the above-mentioned four island countries. Although WHO recently changed recommended ANC visits from ANC4+ to ANC8+, our analyses applied ANC4+ visits because the data used in this paper were collected before the recommendation was made, and the targets for ANC4+ visits and use of SBA are still aiming to reach by 2025 (4, 5). This study has two specific aims. The first aim was to examine the association between the age at first birth and utilization of maternal health services (i.e., use of ANC4+ visits during pregnancy and SBA) in four island countries mentioned above. We hypothesized that older women at first birth would be more likely to have ANC4+ visits and use an SBA than the younger women while controlling for other variables. The second aim was to examine the relationship between maternal health services (i.e., use of ANC4+ and SBA) and demographic characteristics (age, place of residence, education, wealth status, age at first cohabitation; birth orders, number of children), reproductive health knowledge (knowledge of modern contraceptives, ovulation, pregnancy), and household ownership of a vehicle (having a car/truck or a motorcycle/scooter).

The findings from this paper will inform the decision-makers of the current situations and factors that influence the use of maternal health services and will provide appropriate recommendations to reduce maternal mortality in this higher-risk age group.

## Methods

### Data source

Nationally representative data from the Demographic Health Surveys (DHS) of Indonesia (IDHS:2017) (12), the Philippines (PDHS: 2017) (13), Timor-Leste (TLDHS: 2016) (14), and Papua New Guinea (PNGDHS: 2016-2018) (15) were used. For each survey, 49,627 Indonesian women, 25,074 Filipino women, 15,198 Papua New Guinean women, and 12,607 Timor-Leste women participated. The DHS surveys were reviewed and approved by the Institutional Review Board (IRB) of Inner-City fund (ICF) in collaboration with respective country’s Ministry of Health (16). ICF ensures that the DHSs complies with appropriate guidelines and regulations. For DHS conducted in the included countries, written informed consent was obtained from the respondents (16). If the respondents were aged 15-17, parental or guardian consent was taken (12–15).

The outcome variables, having ANC4+ visits and using an SBA at childbirth, were dichotomous. The predictor variables were maternal age at first birth (<18, 19-24, >24), age at first cohabitation (<18, 19-24, >24), age at last birth (<18, 19-24, >24), age groups (<18, 19-24, >24), place of residence (urban and rural), education (no education/primary education, secondary education, higher education), standardized wealth index (poor, middle and rich), birth order (BO) of the children (BO=1, BO=2, BO=3, BO=4 and above) and the number of children (having a child, two children, three children, and four and above). We included three reproductive health-related knowledge variables (knowledge on timing of ovulation, getting pregnant before the first period after giving birth to a baby, and modern contraceptive methods). We also included two transportation indicators: whether the household has a car/truck and if the household has a motorcycle/scooter.

We performed descriptive analyses (weighted prevalence and Chi-square test) to observe the association between the outcome and independent variables. Multivariable analyses (logistic regression) were conducted to observe the relationship between the outcome variables (use of ANC4+visits and SBA) and exposure variable (age at first birth) when controlling for other independent variables. In addition, as we used four age-related variables, we also checked for multicollinearity. All the analyses were applied survey weights and performed using Stata 17.0 (17) (Stata.com, n.d.).

### Study population

For this analysis, the number of respondents who answered the question of ANC were 17,779 from Indonesia, 7,992 from the Philippines, 9,057 from Papua New Guinea, and 7,211 from Timor-Leste. On the other hand, the number of respondents answering the question of SBA were 17,785 from Indonesia, 10,551 from the Philippines, 9,204 from Papua New Guinea, and 7,211 from Timor-Leste, respectively.

## Results

### Descriptive and bivariate analyses

The number of women who had ANC4+ visits ranged from 48% in Papua New Guinea to 90% in Indonesia. The use of SBA ranged from 40% (Indonesia) to 84% (Philippines). The percentages of women giving birth before age 18 ranged from 18% in Indonesia to 31% in Papua New Guinea. Only in Indonesia, bivariate results showed that that all independent variables were significantly related to ANC4+ visits (Table 1).

**Table 1:** Bivariate analyses of having ANC4+ visits (weighted frequency and percentage) in four island countries in Asia and the Pacific region.

| Countries and original observations | Indonesia:<br>49, 627 |  | Philippines:<br>25, 074 |  | Papua New Guinea:<br>15, 198 |  | Timor-Leste:<br>12, 607 |  |
| --- | --- | --- | --- | --- | --- | --- | --- | --- |
| Outcome variables: ANC4+ visits |  |  |  |  |  |  |  |  |
| Sociodemographic Characteristics | No: N (%) | Yes: N (%) | No: N (%) | Yes: N (%) | No: N (%) | Yes: N (%) | No: N (%) | Yes: N (%) |
| Age at first birth |  | 17,779 |  | 7, 992 |  | 9, 057 |  | 7, 211 |
| <19 | 669 (17) | 2,706 (83) | 447 (20) | 1, 491 (80) | 1,314 (54) | 1,228 (46) | 413 (28) | 1, 056 (72) |
| 19-24 | 1, 197 (10) | 8,373 (90) | 640 (13) | 3, 646 (87) | 2,360 (52) | 2,625 (48) | 1,036 (24) | 3, 230 (76) |
| >25 | 421 (7) | 4,408 (94) | 197 (9) | 1, 571 (91) | 686 (48) | 844 (52) | 346 (22) | 1, 130 (78) |
|  |  | P < .001 |  | P < .001 |  | P = .203 |  | P = . 046 |
| Age at first cohabitation |  | 17,750 |  | 7, 683 |  | 8, 768 |  | 7, 179 |
| <19 | 988 (15) | 4, 651 (85) | 608 (18) | 2, 196 (82) | 1, 821 (54) | 1, 741 (46) | 673 (27) | 1, 832 (73) |
| 19-24 | 986 (7) | 7,753 (91) | 481 (11) | 3, 031 (89) | 1, 926 (51) | 2, 196 (49) | 879 (23) | 2, 759 (77) |
| >25 | 306 (7) | 3,066 (93) | 141 (8) | 1, 226 (92) | 473 (49) | 611 (51) | 236 (22) | 800 (78) |
|  |  | P < .001 |  | P < .001 |  | P= .123 |  | P = .013 |
| Age at last birth |  | 16, 998 |  | 7, 992 |  | 8, 518 |  | 6, 851 |
| <19 | 93 (18) | 331 (82) | 136 (23) | 393 (77) | 118 (42) | 134 (58) | 36 (29) | 1, 213 (78) |
| 19-24 | 522 (10) | 3,233 (90) | 366 (13) | 2, 008 (87) | 1, 105 (52) | 1, 243 (48) | 355 (22) | 1, 213 (78) |
| >25 | 1, 574 (10) | 11,245 (90) | 782 (13) | 4, 307 (87) | 2, 867 (52) | 3, 501 (48) | 1, 333 (25) | 3, 841 (75) |
|  |  | P < .001 |  | P < .001 |  | P= .245 |  | P = .105 |
| Age group and SBA |  | 17,779 |  | 7, 992 |  | 9, 507 |  | 7, 211 |
| <19 | 107 (20) | 330 (80) | 83 (21) | 258 (79) | 160 (50) | 146 (50) | 52 (26) | 121 (74) |
| 19-24 | 389 (10) | 2,411 (90) | 298 (16) | 1, 318 (84) | 895 (53) | 1, 004 (4) | 273 (22) | 979 (78) |
| 25 and above | 1, 791 (10) | 12,751 (90) | 903 (13) | 5, 132 (88) | 3, 305 (52) | 3, 547 (48) | 1, 470 (24) | 4, 316 (76) |
|  |  | P < .001 |  | P < .001 |  | P = .888 |  | P = .338 |
| Place of residence |  | 17,779 |  | 7, 992 |  | 9, 057 |  | 7, 211 |
| Urban | 780 (7) | 7,947 (93) | 317 (12) | 2, 285 (88) | 661 (38) | 1, 359 (62) | 347 (13) | 1, 787 (87) |
| Rural | 1, 507 (13) | 7,545 (87) | 967 (15) | 4, 423 (85) | 3, 699 (54) | 3, 338 (46) | 1, 448 (29) | 3, 629 (71) |
|  |  | P < .001 |  | P = .029 |  | P < .001 |  | P < .001 |
| Education |  | 17,779 |  | 7, 992 |  | 9, 057 |  | 7, 211 |
| No education/Primary education | 988 (18) | 3,715 (82) | 517 (28) | 1, 121 (72) | 3, 543 (58) | 2, 900 (42) | 1, 087 (34) | 2, 070 (66) |
| Middle | 1,071 (8) | 8,812 (92) | 575 (14) | 3, 370 (87) | 729 (33) | 1, 547 (67) | 634 (18) | 2, 834 (82) |
| Higher | 228 (4) | 2,965 (96) | 192 (6) | 2, 217 (94) | 88 (37) | 250 (63) | 74 (11) | 512 (89) |
|  |  | P < .001 |  | P < .001 |  | P < .001 |  | P < .001 |
| Wealth |  | 17, 779 |  | 7, 992 |  | 9, 057 |  | 7, 211 |
| Poorest/Poorer | 1, 653 (17) | 6, 758 (83) | 978 (20) | 3, 582 (81) | 1, 934 (63) | 1, 1125 (37) | 1, 055 (36) | 1, 889 (64) |
| Middle | 300 (8) | 2, 947 (92) | 170 (12) | 1, 274 (88) | 858 (50) | 872 (50) | 360 (25) | 1, 105 (75) |
| Richest/Richer | 334 (4) | 5, 787 (96) | 136 (6) | 1, 852 (94) | 1, 568 (41) | 2, 700 (59) | 380 (12) | 2, 422 (88) |
|  |  | P < .001 |  | P < .001 |  | P < .001 |  | P < .001 |
| Knowledge on ovulation |  | 17, 767 |  | 7, 992 |  | 9, 010 |  | 7, 211 |
| No | 1, 925 (11) | 11, 474 (89) | 992 (14) | 4, 782 (86) | 3, 514 (53) | 3, 539 (47) | 1, 626 (25) | 4, 797 (76) |
| Yes | 362 (7) | 4, 006 (93) | 292 (12) | 1, 926 (88) | 824 (48) | 1, 133 (53) | 169 (21) | 619 (79) |
|  |  | P < .001 |  | P = .075 |  | P = .043 |  | P = .176 |
| **Knowledge on getting pregnant** |  | **17, 774** |  | **7, 992** |  | **9, 002** |  | **7, 211** |
| No | 1, 192 (13) | 6, 234 (87) | 294 (16) | 1, 256 (84) | 1, 946 (58) | 1, 705 (43) | 1, 125 (29) | 2, 696 (71) |
| Yes | 1, 095 (9) | 9, 253 (92) | 990 (13) | 5, 452 (87) | 2, 385 (48) | 2, 966 (52) | 670 (19) | 2, 720 (81) |
|  |  | P < .001 |  | P = .053 |  | P < .001 |  | P < .001 |
| **Knowledge on modern contraception** |  | **17, 779** |  | **7, 992** |  | **9, 057** |  | **7, 211** |
| No | 69 (82) | 26 (18) | 26 (80) | 7 (20) | 861 (81) | 219 (19) | 549 (42) | 728 (58) |
| Yes | 2, 218 (10) | 15, 466 (90) | 1, 258 (13) | 6, 701 (87) | 3, 499 (47) | 4, 478 (53) | 1, 246 (20) | 4, 688 (80) |
|  |  | P < .001 | P < .001 |  | P < .001 |  | P < .001 | P < .001 |
| **Birth Order (BO)** |  | **17, 779** |  | **7, 992** |  | **9, 507** |  | **7, 211** |
| BO=1 | 627 (8) | 5, 218 (92) | 283 (11) | 1, 913 (90) | 955 (46) | 1, 299 (54) | 374 (20) | 1, 372 (80) |
| BO=2 | 570 (8) | 5, 018 (92) | 276 (12) | 1, 801 (88) | 870 (51) | 1, 058 (49) | 306 (21) | 1, 120 (79) |
| BO=2 | 407 (11) | 2,943 (90) | 216 (13) | 1, 282 (87) | 810 (53) | 821 (47) | 289 (24) | 915 (76) |
| BO=3 | 683 (22) | 2, 313 (78) | 509 (19) | 1, 712 (81) | 1, 725 (57) | 1, 519 (43) | 826 (29) | 2, 009 (71) |
|  |  | P < .001 |  | P < .001 |  | P < .001 |  | P < .001 |
| **Number of Children** |  | **17, 704** |  | **7, 964** |  | **9, 000** |  | **7, 173** |
| One child | 481 (7) | 4, 581 (93) | 287 (10) | 1, 958 (90) | 656 (42) | 961 (58) | 248 (21) | 856 (79) |
| Two children | 608 (8) | 5, 526 (92) | 280 (12) | 1, 826 (88) | 913 (52) | 1, 157 (49) | 308 (18) | 1, 326 (82) |
| Three children | 495 (12) | 3, 140 (89) | 218 (13) | 1, 275 (87) | 908 (53) | 925 (47) | 317 (23) | 1, 031 (77) |
| Four children | 681 (23) | 2, 192 (77) | 488 (19) | 1, 632 (81) | 1, 849 (56) | 1, 631 (44) | 904 (29) | 2, 183 (71) |
|  |  | P < .001 |  | P < .001 |  | P < .001 |  | P < .001 |
| **Household has a car** |  | **17, 769** |  | **7, 992** |  | **9, 037** |  | **7, 211** |
| No | 2,154 (11) | 13, 382 (89) | 1, 256 (14) | 6, 291 (86) | 4,184 (53) | 4, 310 (47) | 1, 747 (25) | 5, 112 (75) |
| Yes | 130 (4) | 2, 103 (96) | 28 (4) | 417 (96) | 168 (33) | 375 (67) | 48 (14) | 304 (86) |
|  |  | P < .001 |  | P < .001 |  | P < .001 |  | P = .030 |
| **Household has a motorbike** |  | **17, 768** |  | **7, 992** |  | **9, 036** |  | **7, 211** |
| No | 883 (19) | 3, 009 (81) | 930 (16) | 3, 935 (84) | 4, 331 (52) | 4, 4639 (48) | 1, 364 (30) | 3, 063 (70) |
| Yes | 1, 400 (8) | 12, 476 (92) | 354 (9) | 2, 773 (91) | 21 (34) | 45 (66) | 431 (14) | 2, 353 (86) |
|  |  | P < .001 |  | P < .001 |  | P = .078 |  | P < .001 |

Furthermore, there was a significant association between the use of SBA and all the independent variables in all countries, except the variables age at last birth, knowledge on ovulation and getting pregnant in the Philippines and the age groups in Timor-Leste (Table 2).

**Table 2:** Bivariate analyses of the use of SBA (weighted frequency and percentage) in four island countries in Asia and the Pacific region.

| Country & Original observations | Indonesia: 49, 627 | Philippines: 25, 074 | Papua New Guinea: 15, 198 | Timor-Leste: 12, 607 |
| --- | --- | --- | --- | --- |
| Outcome variables: ANC4+ visits |  |  |  |  |

| **Sociodemographic characteristics** | No: N (%) | Yes: N (%) | No: N (%) | Yes: N (%) | No: N (%) | Yes: N (%) | No: N (%) | Yes: N (%) |
| --- | --- | --- | --- | --- | --- | --- | --- | --- |
| **Age at first birth** |  | **17, 785** |  | **10, 551** |  | **9, 204** |  | **7, 211** |
| <19 | 2,541 (74) | 835 (26) | 795 (23) | 1, 895 (77) | 1, 155 (49) | 1,431 (51) | 757 (54) | 712 (46) |
| 19-24 | 6, 302 (64) | 3, 278 (37) | 1, 123 (16) | 4, 518 (85) | 1, 823 (42) | 3, 236 (58) | 1, 783 (43) | 2, 483 (57) |
| >25 | 2, 302 (44) | 2, 527 (56) | 259 (8) | 1, 961 (92) | 520 (38) | 1, 039 (62) | 516 (35) | 960 (65) |
|  |  | P < .001 |  | P < .001 |  | P < .001 |  | P < .001 |
| **Age at first cohabitation** |  | **17, 756** |  | **10, 206** |  | **8, 911** |  | **7, 179** |
| <19 | 3, 139 (72) | 1, 503 (28) | 1, 159 (24) | 2, 703 (77) | 1, 574 (48) | 2, 038 (52) | 1, 237 (52) | 1, 268 9 (48) |
| 19-24 | 5, 438 (59) | 3, 304 (41) | 794 (13) | 3, 841 (87) | 1, 445 (40) | 2, 741 (60) | 1, 453 (40) | 2, 185 (60) |
| >25 | 1, 545 (41) | 1, 827 (59) | 196 (8) | 1, 513 (92) | 388 (42) | 725 (58) | 355 (34) | 681 (66) |
|  |  | P < .001 |  | P < .001 |  | P < .001 |  | P < .001 |
| **Age at last birth** |  | **17, 004** |  | **10, 551** |  | **8, 662** |  | **6, 851** |
| <19 | 327 (76) | 97 (24) | 122 (16) | 476 (84) | 82 (37) | 174 (63) | 45 (38) | 64 (62) |
| 19-24 | 2, 572 (66) | 1, 184 (34) | 642 (15) | 2, 551 (85) | 795 (38) | 1, 583 (62) | 617 (40) | 951 (60) |
| >25 | 7, 760 (58) | 5, 064 (42) | 1, 413 (16) | 5, 347 (84) | 2, 410 (46) | 3, 618 (54) | 2, 251 (44) | 2, 923 (56) |
|  |  | P < .001 |  | P = .528 |  | P < .001 |  | P = .033 |
| **Age group and SBA** |  | **17, 785** |  | **10,551** |  | **9,204** |  | **7,211** |
| <19 | 328 (72) | 109 (28) | 78 (13) | 319 (87) | 96 (39) | 216 (61) | 71 (42) | 102 (58) |
| 19-24 | 1, 926 (66) | 875 (34) | 449 (16) | 1, 767 (84) | 646 (37) | 1, 275 (63) | 482 (39) | 770 (61) |
| 25 and above | 8, 891 (59) | 5, 656 (41) | 1, 650 (16) | 6, 288 (84) | 2, 756 (45) | 4, 215 (55) | 2, 503 (44) | 3, 283 (56) |
|  |  | P < .001 |  | P < .001 |  | P < .001 |  | P = .096 |
| **Place of residence** |  | **17, 785** |  | **10, 551** |  | **9, 204** |  | **7, 211** |
| Urban | 4, 461 (50) | 4, 269 (50) | 318 (8) | 3, 009 (92) | 184 (13) | 1, 869 (87) | 356 (13) | 1, 778 (87) |
| Rural | 6, 684 (70) | 2, 371 (30) | 1, 859 (21) | 5, 365 (79) | 3, 314 (47) | 3, 837 (53) | 2, 700 (55) | 2, 377 (45) |
|  |  | P < .001 |  | P < .001 |  | P < .001 |  | P < .001 |
| **Education** |  | **17, 785** |  | **10, 551** |  | **9, 204** |  | **7, 211** |
| No education/Primary education | 3, 661 (76) | 1, 043 (24) | 1, 119 (41) | 1, 269 (59) | 3, 151 (53) | 3, 412 (48) | 1, 904 (62) | 1, 253 (38) |
| Middle | 6, 148 (59) | 3, 740 (41) | 886 (13) | 4, 319 (87) | 330 (18) | 1, 968 (82) | 1, 115 (33) | 2, 353 (67) |
| Higher | 1, 336 (36) | 1, 857 (64) | 172 (3) | 2, 786 (97) | 17 (3) | 326 (97) | 37 (5) | 549 (95) |
|  |  | P < .001 |  | P < .001 |  | P < .001 |  | P < .001 |
| **Wealth** |  | **17, 785** |  | **10, 551** |  | **9, 204** |  | **7, 211** |
| Poorest/Poorer | 6, 498 (75) | 1, 915 (25) | 1, 985 (27) | 4, 393 (73) | 2, 009 (65) | 1,096 (35) | 1, 907 (67) | 1, 037 (33) |
| Middle | 1, 955 (61) | 1, 293 (39) | 134 (7) | 1, 673 (93) | 735 (44) | 1, 020 (56) | 620 (44) | 845 (56) |
| Richest/Richer | 2, 692 (44) | 3, 432 (56) | 58 (2) | 2, 308 (98) | 754 (20) | 3, 590 (80) | 529 (18) | 2, 273 (82) |
|  |  | P < .001 |  | P < .001 |  | P < .001 |  | P < .001 |
| **Knowledge on ovulation** |  | **17, 773** |  | **10, 551** |  | **9, 152** |  | **7, 211** |
| No | 8, 797 (63) | 4, 606 (37) | 1, 621 (16) | 6, 000 (84) | 2, 875 (46) | 4, 278 (54) | 2, 797 (45) | 3, 626 (56) |
| Yes | 2, 342 (50) | 2, 028 (50) | 556 (14) | 2, 374 (86) | 606 (36) | 1, 393 (64) | 259 (32) | 529 (68) |
|  |  | P < .001 |  | P = 0.177 |  | P < .001 |  | P < .001 |
| **Knowledge on getting pregnant** | **17, 780** |  | **10, 551** |  | **9, 146** |  | **7, 211** |  |
| No | 4,824 (62) | 2, 602 (38) | 407 (15) | 1, 633 (85) | 1, 572 (48) | 2, 138 (52) | 1, 686 (46) | 2, 135 (54) |
| Yes | 6,320 (59) | 4, 034 (41) | 1, 770 (16) | 6, 741 (84) | 1, 907 (40) | 3, 529 (60) | 1, 370 (41) | 2, 020 (59) |
|  |  | P = .012 |  | P = .657 |  | P < .001 |  | P = .007 |
| **Knowledge on modern contraception** |  | **17, 785** |  | **10, 551** |  | **9, 204** |  | **7, 211** |
| No | 93 (97) | 2 (3) | 36 (70) | 12 (30) | 805 (73) | 286 (27) | 738 (59) | 539 (41) |
| Yes | 11, 025 (60) | 6, 638 (40) | 2, 141 (16) | 8, 362 (84) | 2, 693 (39) | 5, 420 (61) | 2, 318 (40) | 3, 616 (60) |
|  |  | P < .001 |  | P < .001 |  | P < .001 |  | P < .001 |
| **Birth Order (BO)** |  | **17,785** |  | **10,551** |  | **9,204** |  | **7,211** |
| BO=1 | 3, 383 (56) | 2, 463 (44) | 388 (9) | 2, 706 (91) | 616 (31) | 1, 663 (69) | 523 (30) | 1, 223 (70) |
| BO=2 | 3, 429 (59) | 2, 161 (41) | 447 (12) | 2, 255 (88) | 645 (38) | 1, 314 (62) | 567 (42) | 859 (58) |
| BO=2 | 2, 145 (62) | 1, 206 (38) | 384 (15) | 1, 480 (85) | 661 (47) | 999 (53) | 532 (45) | 672 (54) |
| BO=3 | 2, 188 (71) | 810 (29) | 958 (28) | 1, 933 (72) | 1, 576 (53) | 1, 730 (47) | 1, 434 (51) | 1, 401 (49) |
|  |  | P < .001 |  | P < .001 |  | P < .001 |  | P < .001 |
| **Number of Children** |  | **17,710** |  | **10,521** |  | **9,147** |  | **7,173** |
| One child | 2, 898 (56) | 2, 165 (44) | 247 (8) | 2, 039 (92) | 448 (31) | 1, 186 (69) | 309 (27) | 795 (73) |
| Two children | 3, 717 (58) | 2, 419 (42) | 426 (11) | 2, 480 (89) | 630 (36) | 1, 475 (64) | 580 (37) | 1, 054 (63) |
| Three children | 2, 355 (62) | 1, 283 (38) | 438 (16) | 1, 719 (85) | 721 (44) | 1, 137 (56) | 582 (46) | 766 (54) |
| Four children | 2, 128 (72) | 745 (28) | 1, 061 (28) | 2, 111 (72) | 1, 676 (53) | 1, 874 (47) | 1, 572 (52) | 1, 515 (48) |
|  |  | P < .001 |  | P < .001 |  | P < .001 |  | P < .001 |
| **Household has a car** |  | **17, 775** |  | **10, 551** |  | **9, 182** |  | **7, 211** |
| No | 10, 249 (64) | 5, 293 (36) | 2, 162 (17) | 7, 859 (84) | 3, 452 (45) | 5, 174 (55) | 3, 010 (45) | 3, 849 (55) |
| Yes | 892 (38) | 1, 341 (62) | 15 (3) | 515 (97) | 38 (8) | 518 (92) | 46 (12) | 306 (88) |
|  |  | P < .001 |  | P < .001 |  | P < .001 |  | P < .001 |
| **Household has a motorbike** |  | **17, 774** |  | **10, 551** |  | **9, 181** |  | **7, 211** |
| No | 2, 976 (73) | 917 (27) | 1, 681 (19) | 4, 963 (81) | 3, 486 (44) | 5, 628 (56) | 2, 367 (54) | 2, 060 (46) |
| Yes | 8, 165 (58) | 5, 716 (43) | 496 (10) | 3, 411 (90) | 3 (2) | 64 (98) | 689 (25) | 2, 095 (75) |
|  |  | P < .001 |  | P < .001 |  | P < .001 |  | P < .001 |

### Multivariable logistic regression results: factors influencing having ANC4+ visits

We presented the detailed results of the logistic regression models for having ANC4+ visits in Table 3. We observed no significant association between ANC4+ visits and age at first birth or age at first cohabitation in all countries. Having ANC4+ visits showed significant association with the age at last birth only in the Philippines and Papua New Guinea. In the Philippines, the women whose age at last birth at older than 19 were more likely to use ANC4+ visits (adjusted odds ratio (aOR)= 1.94; 95% CI=1.23-3.05 among women group aged 19-24) than those whose age at last birth younger than 19. On contrary, a significant relationship was seen in Papua New Guinean women of aged at last birth 19-24 were less likely to use ANC4+ visits than those younger than 19 years (aOR= 0.48; 95% CI=0.29-0.80).

**Table 3:** Multivariable results: Relationship between ANC4+ and sociodemographic characteristics in four island countries in Asia and the Pacific region.

| Country name and number of observations | Indonesia: 16,870 | Philippines: 7,660 | Papua New Guinea: 8,116 | Timor-Leste: 6,784 |
| --- | --- | --- | --- | --- |
| Sociodemographic characteristics | aOR (95% CI) | aOR (95% CI) | aOR (95% CI) | aOR (95% CI) |
| **Age at first birth** |  |  |  |  |
| <19 (Reference) |  |  |  |  |
| 19-24 | 1.17 (0.92-1.49) | 0.85 (0.65-1.12) | 0.92 (0.73-1.17) | 0.97 (0.72-1.30) |
| >25 | 1.19 (0.79-1.81) | 0.70 (0.43-1.16) | 0.87 (0.62-1.22) | 0.94 (0.61-1.46) |
| **Age at first cohabitation** |  |  |  |  |
| <19 (Reference) |  |  |  |  |
| 19-24 | 1.02 (0.81-1.27) | 1.13 (0.86-1.50) | 0.94 (0.76-1.16) | 1.05 (0.80-1.39) |
| >25 | 0.90 (0.59-1.37) | 1.24 (0.74-2.07) | 1.00 (0.71-1.40) | 1.05 (0.69-1.60) |
| **Age at last birth** |  |  |  |  |
| <19 (Reference) |  |  |  |  |
| 19-24 | 1.23 (0.73-2.07) | 1.94 (1.23-3.05) ** | 0.48 (0.29-0.80) ** | 1.50 (0.61-3.69) |
| >25 | 1.81 (0.94-3.48) | 1.85 (1.02-3.37) * | 0.62 (0.34-1.11) | 1.52 (0.56-4.11) |
| **Age group and ANC** |  |  |  |  |
| <19 (Reference) |  |  |  |  |
| 19-24 | 1.59 (0.94-2.69) | 0.82 (0.51-1.31) | 1.56 (0.93-2.63) | 0.92(0.43-1.97) |
| 25 and above | 1.83 (0.99-3.38) * | 1.47 (0.82-2.62) | 1.66 (0.93-2.95) | 0.87 (0.38-2.03) |
| **Place of residence** |  |  |  |  |
| Urban (Reference) |  |  |  |  |
| Rural | 0.83 (0.69-1.00) * | 1.04 (0.76-1.41) | 0.84 (0.59-1.18) | 0.79(0.56-1.11) |
| **Education** |  |  |  |  |
| No education/Primary education (Reference) |  |  |  |  |
| Middle | 1.47 (1.23-1.75) *** | 1.93 (1.51-2.46) *** | 2.06 (1.67-2.54) *** | 1.54 (1.24-1.90) *** |
| Higher | 1.58 (1.15-2.17) ** | 3.11 (2.16-4.47) *** | 1.73 (0.79-3.77) | 1.41(0.84-2.38) |
| **Wealth** |  |  |  |  |
| Poorest/Poorer (Reference) |  |  |  |  |
| Middle | 1.64 (1.30-2.06) *** | 1.30 (0.96-1.77) | 1.41 (1.12-1.78) ** | 1.38 (1.09-1.76) ** |
| Richest/Richer | 2.39 (1.84-3.11) *** | 1.74 (1.18-2.57) ** | 1.55 (1.16-2.07) ** | 2.45 (1.78- 3.35) *** |
| **Knowledge on ovulation** |  |  |  |  |
| No (Reference) |  |  |  |  |
| Yes | 1.25 (1.03-1.52) * | 1.04 (0.83 -1.30) | 1.06 (0.83-1.36) | 0.87(0.62-1.24) |
| **Knowledge on getting pregnant** |  |  |  |  |
| No (Reference) |  |  |  |  |
| Yes | 1.32 (1.15-1.52) *** | 1.31 (1.00 -1.71) * | 1.20(1.02-1.42) * | 1.48 (1.21-1.81) *** |
| **Knowledge on modern contraception** |  |  |  |  |
| No (Reference) |  |  |  |  |
| Yes | 12.32 (4.82-31.49) *** | 18.40 (6.45-52.52) *** | 3.18 (2.22-4.54) *** | 2.20 (1.74-2.77) *** |
| **Birth Order (BO)** |  |  |  |  |
| BO=1 (Reference) |  |  |  |  |
| BO=2 | 1.43 (1.13-1.81) ** | 0.90(0.40-2.01) | 1.06 (0.87-1.29) | 0.95(0.79-1.16) |
| BO=3 | 2.15 (1.49-3.11) *** | 0.76 (0.27-2.14) | 0.93 (0.68 -1.28) | 1.05 (0.77-1.44) |
| BO=4 | 2.12 (1.35-3.34) *** | 0.55 (0.16-1.84) | 0.88 (0.61 -1.27) | 1.12 (0.79-1.57) |
| **Number of Children** |  |  |  |  |
| One child (Reference) |  |  |  |  |
| Two children | 0.48 (0.35-0.66) *** | 0.69 (0.31-1.57) | 0.61 (0.45-0.85) ** | 1.20 (0.87-1.66) |
| Three children | 0.21 (0.13-0.32) *** | 0.67 (0.25-1.79) | 0.62 (0.40-0.96) * | 0.94 (0.65-1.37) |
| Four children | 0.10 (0.06-0.18) *** | 0.70 (0.21-2.30) | 0.56 (0.35-0.90) ** | 0.71 (0.46-1.10) |
| **Household has a car** |  |  |  |  |
| No (Reference) |  |  |  |  |
| Yes | 1.28 (0.94-1.74) | 1.91 (1.06-3.44) * | 0.98(0.65-1.49) | 0.69 (0.37 -1.27) |
| **Household has a motorbike** |  |  |  |  |
| No (Reference) |  |  |  |  |
| Yes | 1.36 (1.13-1.63) *** | 1.31 (1.04 -1.66) * | 1.35 (0.51 -3.52) | 1.26 (0.99-1.61) |
| **Constant** | 0.17 (0.06-0.46) *** | 0.10 (0.03-0.31) *** | 0.45 (0.20-0.98) * | 0.77 (0.41-1.46) |

Note: \_cons estimate baseline odds. P-value: \*p < .05. \*\*p < .01. \*\*\*p < .001; aOR= adjusted Odds Ratio; 95% CI= 95% Confidence Interval

Wealth status, having knowledge of getting pregnant, and having knowledge of modern contraception significantly predicted the use of ANC4+ visits. Wealthier women were more likely to use ANC4+ visits than those who were poor, and the highest odds ratio was seen in richer/richest women in Indonesia (aOR=2.39; 95% CI=1.84-3.11).

The women who had knowledge about getting pregnant were more likely to use ANC4+ visits than those who did not in all four countries. The highest odds ratio was seen in Timor-Leste (aOR= 1.48, 95% CI=1.21-1.81). Similarly, the use of ANC4+ visits showed significant associations with having knowledge of the modern contraception in all four countries; odds ratio being the highest in the Philippines (aOR= 18.40; 95% CI= 6.45 -52.52).

The variables such as education, place of residence, knowledge on ovulation, household ownership of a vehicle, birth order, and number of children showed different patterns of ANC4+ visits in four countries. The highest odds ratio was observed among women with a higher education than those without or those with primary education in Papua New Guinea (aOR=2.06, 95% CI= 1.67-2.54). The rural women in Indonesia were less likely to have ANC4+ visits than urban women (aOR=0.83, 95% CI= 0.69-1.00). However, there were no significant associations between the use of ANC4+ visits and place of residence in the Philippines, Papua New Guinea and Timor-Leste. Similarly, only Indonesia women who had knowledge on ovulation were more likely to use ANC4+ visits than those who did not have such knowledge (aOR=1.25, 95% CI= 1.03-1.52). The association between ANC4+ visits and other factors such as birth order, number of children, reproductive knowledge, and household ownership of a vehicle showed very different patterns in four countries (Table 3).

The association between the use of ANC4+ visits and household ownership of a car/truck was significant only in the Philippines (aOR=1.91; 95% CI=1.06-3.44), and household ownership of a motorbike was significant in Indonesia (aOR=1.36, 95% CI= 1.13-1.63) and the Philippines (aOR=1.31: 95% CI= 1.35 (0.51-3.52).

### Multivariable logistic regression results: factors influencing the use of SBA

Table 4 shows the result of logistic regression models for the use of SBA. There was no significant association between utilization of SBA and the age at first birth in all countries except in Indonesia. In Indonesia, the women aged 25 and older were 1.38 times (95% CI=1.09-1.74) more likely to use SBA than the women who were younger than 19 and the women who age at first cohabitation at aged 25 and older were 1.45 times (95% CI=1.18-1.79) more likely to use SBA than those who were younger than aged 18. Similar pattern was observed for the women with age at last birth 25 and older. They were 1.67 times (95% CI= 1.03-2.71) more likely to use SBA than the younger women aged under 19. In the Philippines, the women older than 19 at age at last birth were more likely to use SBA than those who are under 19 and the association was significant. Other two countries did not show any significant association between the use of SBA and age-related variables.

**Table 4:** Multivariable results: Relationship between use of SBA and sociodemographic characteristics in four island countries in Asia and the Pacific region.

| Country and Number of observations | Indonesia: 16,876 | Philippines: 10,181 | Papua New Guinea: 8,249 | Timor-Leste: 6,784 |
| --- | --- | --- | --- | --- |
| Sociodemographic characteristics | aOR (95% CI) | aOR (95% CI) | aOR (95% CI) | aOR (95% CI) |
| **Age at first birth** |  |  |  |  |
| <19 (Reference) |  |  |  |  |
| 19-24 | 1.06 (0.89-1.25) | 0.87 (0.69-1.09) | 0.93 (0 .72-1.20) | 1.07 (0.81-1.40) |
| >25 | 1.38 (1.09-1.74) ** | 1.04 (0.64-1.69) | 1.17(0.85-1.61) | 1.24 (0.80-1.92) |
| **Age at first cohabitation** |  |  |  |  |
| <19 (Reference) |  |  |  |  |
| 19-24 | 1.12 (0.97-1.28) | 1.23 (0.98-1.53) | 1.16 (0.92-1.47) | 1.18 (0.95-1.47) |
| >25 | 1.45 (1.18-1.79) *** | 1.04 (0.67-1.62) | 0.92 (0.66-1.28) | 1.23 (0.82-1.84) |
| **Age at last birth** |  |  |  |  |
| <19 (Reference) |  |  |  |  |
| 19-24 | 1.31 (0.84-2.04) | 1.77 (1.11- 2.84) ** | 0.74(0.44-1.27) | 0 .63 (0.25-1.58) |
| >25 | 1.67 (1.03-2.71) * | 2.31 (1.23-4.33) ** | 0.59 (0.31-1.15) | 0.62 (0.23-1.74) |
| **Age group and SBA** |  |  |  |  |
| <19 (Reference) |  |  |  |  |
| 19-24 | 0.92(0.62-1.38) | 0.47(0.28-0.79) ** | 1.17 (0.68-2.00) | 1.33 (0 .55 -3.21) |
| 25 and above | 0.81(0.53-1.26) | 0 .49 (0.25-0.95) * | 1.34 (0.68-2.62) | 1.18 (0.46-2.99) |
| **Place of residence** |  |  |  |  |
| Urban (Reference) |  |  |  |  |
| Rural | 0.66 (0.58-0 .74) *** | 0 .55 (0.38-0.81) ** | 0 .49 (0.25-0.94) ** | 0.32(0.25-0.42) *** |
| **Education** |  |  |  |  |
| No education/Primary education (Reference) |  |  |  |  |
| Middle | 1.43 (1.27-1.62) *** | 2.73 (2.10-3.54) *** | 2.81 (2.15-3.68) *** | 1.63 (1.35-1.97) *** |
| Higher | 2.11 (1.79-2.48) *** | 6.02 (4.09-8.87) *** | 8.46 (3.93-18.23) *** | 5.29 (3.25-8.61) *** |
| **Wealth** |  |  |  |  |
| Poorest/Poorer (Reference) |  |  |  |  |
| Middle | 1.45 (1.26-1.66) *** | 2.49 (1.80-3.46) *** | 1.98 (1.60-2.44) *** | 2.03 (1.59 - 2.58) *** |
| Richest/Richer | 1.92 (1.66-2.21) *** | 6.07 (3.30 -11.14) *** | 3.62 (2.84 -4.63) *** | 3.50 (2.66-4.62) *** |
| **Knowledge on ovulation** |  |  |  |  |
| No (Reference) |  |  |  |  |
| Yes | 1.21 (1.09-1.34) *** | 0.97 (0.78 -1.21) | 1.15 (0.92-1.44) | 1.19 (0 .88-1.61) |
| **Knowledge on getting pregnant** |  |  |  |  |
| No (Reference) |  |  |  |  |
| Yes | 0.93 (0.85-1.02) | 0.93(0.71-1.23) | 1.08 (0.90-1.29) | 0.78 (0.66-0.92) ** |
| **Knowledge on modern contraception** |  |  |  |  |
| No (Reference) |  |  |  |  |
| Yes | 8.08 (2.16-30.24) ** | 5.67 (1.59-20.24) ** | 2.27 (1.63-3.16) *** | 1.62 (1.30-2.02) *** |
| **Birth Order (BO)** |  |  |  |  |
| BO=1 (Reference) |  |  |  |  |
| BO=2 | 1.07 (0.91-1.26) | 0.86 (0.65-1.14) | 0.75(0.60-0.95) ** | 0 .72(0.58-0 .89) ** |
| BO=3 | 1.38 (1.09- 1.74) ** | 1.07 (0.77 -1.49) | 0.55(0.38-0.78) *** | 0 .85 (0.61-1.18) |
| BO=4 | 1.73 (1.28-2.34) *** | 0.82 (0.50-1.34) | 0.58 (0.39-0 .87) ** | 0 .90 (0.61 -1.34) |
| **Number of Children** |  |  |  |  |
| One child (Reference) |  |  |  |  |
| Two children | 0 .80 (0.67-0.95) ** | 0.85 (0.58-1.23) | 1.09 (0.76-1.55) | 0 .75(0.56- 0.99) * |
| Three children | 0.59 (0.46-0 .75) *** | 0.55 (0.36-0.84) ** | 1.01 (0.66-1.54) | 0.58 (9.40-0.85) ** |
| Four children | 0 .36 (0.26-0.50) *** | 0.42 (0.25- 0.72) ** | 0 .91 (0.57- 1.44) | 0.49 (0.30-0.79) ** |
| **Household has a car** |  |  |  |  |
| No (Reference) |  |  |  |  |
| Yes | 1.43 (1.25-1.65) *** | 0.80 (0 .32-2.00) | 1.99 (1.02-3.87) * | 1.37 (0 .83-2.25) |
| **Household has a motorbike** |  |  |  |  |
| No (Reference) |  |  |  |  |
| Yes | 1.17 (1.03-1.33) ** | 1.11 (0.86-1.42) | 8.99 (1.96-41.29) ** | 1.21 (0.98-1.49) |
| Constant | 0.03 (.01- 0.12) *** | 0.72 (0.19-2.81) | 0.88 (0.34-2.25) | 1.75 (0.99-3.08) * |

Note: \_cons estimate baseline odds. P-value: \*p < .05. \*\*p < .01. \*\*\*p < .001; aOR= adjusted Odds Ratio; 95% CI= 95% Confidence Interval

Place of residence, education, wealth status, and having knowledge on modern contraceptives were significant predictors for SBA use in all four countries. The women in rural areas were less likely to use SBA than those living in the urban, and the association was significant in all four countries. The more the women have education, the more likelihood of using SBA than those without education or only having primary education. We observed the highest odds ratio (aOR=8.46, 95% CI=3.93-18.23) in Papua New Guinea and the lowest odds ratio in Indonesia (aOR=2.11, 95% CI= 1.79-2.48) among women with higher education than those who did not have any education or only have primary education. Our results also showed that women in the rich group were almost two times (aOR=1.92, 95% CI= 1.66-2.21 in Indonesia) to six times (aOR=6.07, 95% CI= 3.30-11.14 in the Philippines) more likely to use SBA than the women in the poor group. Women who had knowledge of modern contraceptives were more likely to use SBA than those who did not. The highest odds ratio was seen in Indonesia (aOR=8.08, 95% CI= 2.16-30.24), and the lowest odds ratio was seen in Timor-Leste (aOR=1.62, 95% CI= 1.30-2.02).

The association between birth order and a woman’s use of SBA and between number of children showed very different pattern in four countries. For example, we observed that the women were less likely to use of SBA for their birth order at two in Timor-Leste (aOR= 0 .72, 95% CI= 0.58-0 .89). Similarly, the women were less likely to use SBA if they have two children and more in Indonesia and Timor-Leste, and three children in the Philippines, and the association showed significant (Table 4).

The association between the use of SBA and two reproductive knowledge-related questions (knowledge on getting pregnant, and knowledge on ovulation), and household ownership of a vehicle showed different patterns in these four countries. The women who had knowledge on ovulation were more likely to use SBA than those who did not have, and the association was significant only in Indonesia (aOR= 1.21, 95% CI: 1.09-1.34). The women who had knowledge on getting pregnant after birth before the first period were less likely to use SBA than those who did not have, and the association was significant only in Timor-Leste (aOR=0.78, 95% CI=0.66-0.92). Women who had a car or motorcycle were more likely to use SBA than those who did not have, and the association was significant only in Indonesia and Papau New Guinea (Table 4).

We also tested different models to see if the inclusion of four age-related variables has multicollinearity effect. Since there is no significant increment in standard error (SE) of tested models, we assumed no multicollinearity in our models.

## Discussion

The primary purpose of this study was to analyze the associations between the age at first birth and utilization of two main outcome variables: (a) use of ANC4+ visits during pregnancy and (b) SBA at the time of delivery. We also examined the relationship between social determinants and two main outcome variables. Compared to other high and upper-middle income countries in Asia and the Pacific region, where almost all births used SBA, this finding highlighted an urgent need to find out the reasons the women did not use SBA, the quality of health information given during the ANC visits, or other influencing factors including cultural practices in all these four countries (Indonesia, the Philippines, Papua New Guinea, and Timor-Leste) to reduce the maternal deaths.

### Having antenatal care four or more (ANC4+) visits

The low percentage of women having ANC4+ visits especially in Papua New Guinea (48%) indicated that there is ample room to enhance pregnancy-related health outcomes. A study conducted in Papua New Guinea found several barriers to regular antenatal care: distance to health facility, cost, quality of care, and women’s limited knowledge on the benefit of ANC visits (18).

In 2016, WHO recommended ANC visits increased from ANC4+ to ANC8+. And therefore, we also examined the data on ANC8+ visits (4). However, even with ANC4+ visits, women in these countries faced a lot of barriers. If we want to achieve the new recommended ANC visit coverage target (i.e., 90% of pregnant women to have ANC8+ visits by 2030), the respective governments will have to put forth extra effort (4).

Findings of no significant association of having ANC4+ visits among women who had their first age at birth in all four countries is consistent with mixed findings from different studies (18–20). Our findings on the likelihood of using ANC4+ visits were significantly lower among women who started childbearing at a younger age than those who started at an older age was contrary to a study in Nigeria, where older women were less likely to have ANC visits, but not in Ethiopia, suggesting that different underlying mechanisms drives the women’s ANC utilization (19). The culture of decision-making of the husband or elders may have influenced the younger women’s decision on health care use, which reminds us that future interventions may benefit by including family members (9, 18, 20).

The significant association between the use of ANC4+ visits and factors such as having higher education and being wealthier were consistent with previous studies conducted in Senegal and Nigeria, suggesting that there was a huge health disparity on accessibility to care in these island countries (6, 19). Our results on the greater likelihood of using ANC4+ visits among the women who had reproductive health knowledge was a new finding. In the ANC4+ visits model, talking about contraception was covered at the third visit; perhaps providing reproductive health knowledge to any women of childbearing age could be beneficial (7).

Women at their first pregnancy, especially in Indonesia, were less likely to use ANC4+ visits than those with subsequent pregnancies highlighted a need to find strategies to reach unigravida women, who face greater life-threatening risks in pregnancy (6). The significant association between a vehicle ownership and use of ANC4+ visits in a couple of countries such as the Philippines and Indonesia illustrated a need to explore alternative strategies for pregnant women who did not have any vehicles.

### Use of skilled birth attendants (SBA)

Compared to women’s utilization on ANC4+ visits, the use of SBA was low in all countries ranging from 40% in Indonesia to 84% in the Philippines, signaling health systems in the need for those countries to put forth extra effort to reach 90% SBA coverage. A significant association between SBA use at the time of delivery and the age at first birth was observed only among Indonesian women aged 25 and older, highlighting a need to have more information.

The findings of a positive association between the use of SBA and social determinants such as living in urban areas, having higher education, and being wealthier in all four countries were consistent with the findings from a previous study in low to middle-income countries in other regions suggesting a need to strategies to reach their counterparts (21). Similarly, the fact that the women who have two or more children in Indonesia, the Philippines, and Timor-Leste were less likely to use SBA was consistent with similar findings from a study conducted in Tanzania which also suggested that there is a need to educate to women that every pregnancy is at risk and need to have health care services (22).

A novel finding related to women who had knowledge of modern contraception were more likely to utilize SBA at the time of delivery in all four countries suggests that providing such information to every potential pregnant woman could benefit for their own health although further studies might be useful. Reasons women did not use the SBA at the time of delivery could be multiple: lack of maternal and child health care providers or services in rural areas or providers’ gender or availability of health insurance, trust in Traditional Birth Attendants and transportation (20, 23, 24). Although DHS data we used did not collect variables related to gender or health insurance, we expected that highlighting these variables as potential barriers could increase the decision makers’ awareness on these regards in their policy setting or program implementation, and we discussed more details in the following paragraphs. In 2017-2018, the three countries except Papua New Guinea exceeded the WHO recommendation of 23 per 10,000 population, who were present mostly in the urban cities, showing a huge urban/rural disparity for maternal health services (8). See more the detail of skilled health care workers versus population ratio in those countries in the Table 5.

**Table 5:**
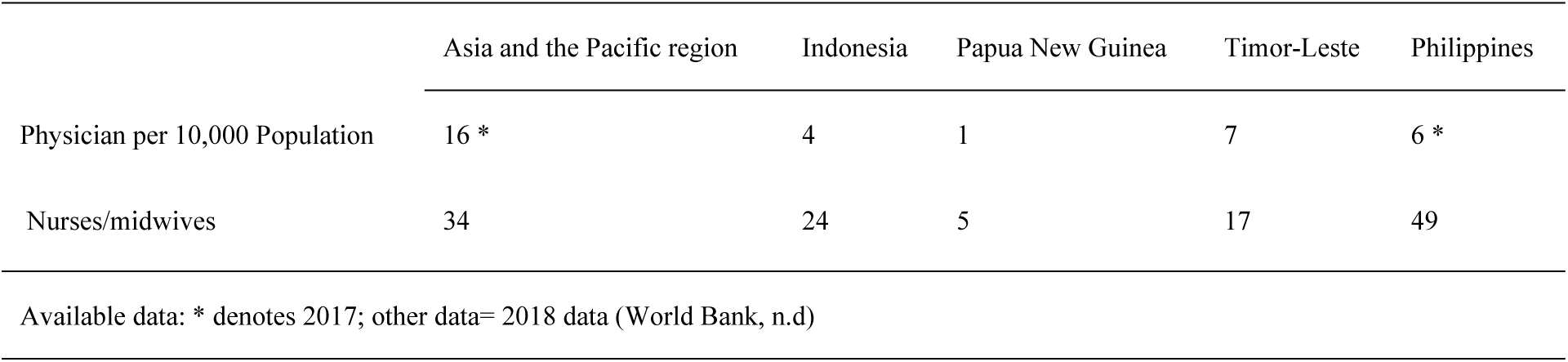
Skilled Health Care workers per 10,000 population in the Asia and the Pacific region and four island countries (2017-2018)

|  | Asia and the Pacific region | Indonesia | Papua New Guinea | Timor-Leste | Philippines |
| --- | --- | --- | --- | --- | --- |
| Physician per 10,000 Population | 16 * | 4 | 1 | 7 | 6 * |
| Nurses/midwives | 34 | 24 | 5 | 17 | 49 |

Available data: \* denotes 2017; other data= 2018 data (World Bank, n.d)

The gender of the SBA could be another influencing factor. A study found that women in PG were reluctant to go to health facilities to give birth because the midwife was a male staff member (25). This information pointed out a need to explore the relationship between gender of SBA and the use of SBA by pregnant women in all these countries.

Health insurance in these countries may also play an influential role in using ANC services and SBA (13, 14, 26–28). In 2018 in Indonesia, only 77% of the population was covered by health insurance program (*Jaminan Kesehatan Nasional-* JKN) which is mostly for curative services (28). In the Philippines, the government established health insurance (Philhealth) covered almost 100% of the population (about 108 million members) in 2019 (13). In Timor-Leste, all the health services are free (27). In the Philippines, health insurance is voluntary and private, giving access only to the wealthier people (26).

Studies in Indonesia and Philippines found that pregnant women in some parts of those countries were more likely to use traditional birth attendants rather than SBA, which could be one of the factors in the limited use of SBA in the rural community (23).

The association between the use of SBA at the time of delivery and the other variables (such as age at first cohabitation, age at last birth, age groups, birth order, household ownership of a car/truck, and a motorcycle/scooter) showed a different pattern of results in these countries. The findings on these different patterns were consistent with a previous study (29). Country specific interventions may be worthwhile to address different age groups’ use of SBA (29).

The mixed findings on association between the birth order and use of SBA in Papua New Guinea, Indonesia, and Timor-Leste highlight a need to explore more in this regard although a previous study from Nigeria found that the women used SBA in their first to six births compared to those who had more than seven births (21). Association between knowledge on getting pregnant and use of SBA showed significant only in Timor-Leste. Also, there were significant association that Indonesian women having knowledge on ovulation were more likely to use SBA than those who did not have. To our knowledge, these two facts were new findings. Our result regarding the association between having a vehicle and use of SBA in Indonesia and Papua New Guinea was consistent with findings from a previous study, in which women with transportation difficulty were less likely to use SBA than those who did not, although their study used road condition as a proxy indicator for transportation barrier (20).

Although our data did not have any traditional and cultural related information, we could not ignore the importance of their roles. Tradition and culturally accepted practices play an influential role in using MCH services, with regional variation within each country (20, 23, 25). In Timor-Leste, decision-making by elders from the family and cultural attitudes towards reproductive health information and services could either be a facilitator or barrier of women’s access to MCH services (20). In Papua New Guinea, transportation was a barrier mentioned by the women as they worried childbirth happened in a vehicle (25, 30). There, the cultural beliefs among some people such as blood from childbirth could poison men and have a polluting effect, etc., which made it difficult to have a vehicle to go to the health facilities and subsequent usage of SBA (25, 30).

### Strengths and limitations

The strength of this study is the utilization of four nationally representative samples. To our knowledge, this is the first study focusing exclusively on island countries in the Asia and Pacific region. However, a causal relationship between the outcome variables and independent variables cannot be established as this is a cross-sectional study.

## Conclusions

The results from this study demonstrated that education, wealth status, knowledge on modern contraception, and place of residence were significant predictors for having ANC4+ visits and use of SBA in all four countries. Our findings regarding the significant association between knowledge on modern contraception and the use of ANC visits and SBA are novel findings that had not been previously examined in these island countries. Providing such knowledge for the adolescents in these countries could enhance their knowledge and use of MCH services as recommended. Also, the age at first birth was significantly associated with the use of ANC visits in Papua New Guinea, ensuring the use of MCH services at a young age at first birth could also help younger women to recognize pregnancy-related life-threatening risk and further prevent maternal morbidity and mortality.

## Data Availability

The data underlying the results presented in the study are available from the Demographic and Health Survey Program (https://dhsprogram.com/data/dataset_admin/login_main.cfmjsessionid=CBF95CCEF6A628F3A8508D8265850D9D.cfusion?CFID=153435621&CFTOKEN=7aad73a421f625a4-4B7BB4AF-E439-9A86-4C4A823800C3E8D6). The person who are going to access the data needs to register and request the data.

